# Impact of the use of cannabis as a medicine in pregnancy, on the unborn child: a systematic scoping review protocol

**DOI:** 10.1101/2024.05.14.24306797

**Authors:** Alexa Ulana Annette Dinant, Yvonne Ann Bonomo, Rachel Canaway, Christine Mary Hallinan

**Affiliations:** Department of Medicine, Faculty of Medicine, Dentistry and Health Sciences, The University of Melbourne, Parkville, VIC 3010, Australia; Department of Addiction Medicine, St. Vincent’s Hospital, Fitzroy, VIC 3065, Australia; Health and Biomedical Research Information Technology Unit (HaBIC R^2^), Department of General Practice and Primary Care, Faculty of Medicine, Dentistry and Health Sciences, The University of Melbourne, Parkville, VIC 3010, Australia

## Abstract

**Introduction:** The use of cannabis for medicinal purposes is on the rise. As more people place their trust in the safety of prescribed alternative plant-based medicine and find it easily accessible, there is a growing concern that pregnant women may be increasingly using cannabis for medicinal purposes to manage their pregnancy symptoms and other health conditions. The objectives of this scoping review are to: conduct a systematic search of the literature to investigate the use of cannabis as a medicine in the context of the recent legislative changes and the resulting increase in use and acceptance, specifically in pregnancy; describe the characteristics of the demographic population using cannabis for medicinal purposes during pregnancy; and to map evidence of its impact on the unborn child and on the child up to twelve months postpartum.

**Methods and analyses:** Research on pregnant women who use cannabis for medicinal purposes only, and infants up to one year after birth who experienced in utero exposure to cannabis for medicinal purposes, will be included in this review. Reviews, randomised controlled trials, case-control, cross-sectional and cohort studies, that have been peer reviewed and published between 1996 and April 2024 as a research paper that investigates prenatal use of cannabis for medicinal purposes and foetal, perinatal, and neonatal outcomes, will be selected for review. Excluding cover editorials, letters, commentaries, protocols, conference papers and book chapters. Effects of illicit drugs use, alcohol misuse and nicotine exposure on neonate outcome will be controlled by excluding studies reporting on the concomitant use of such substances when cannabis data cannot be isolated.

All titles and abstracts will be reviewed by one researcher. Records will be excluded based on title and abstract screening as well as publication type. The full text articles will then be reviewed independently by at least two researchers. Where initial disagreement exists between reviewers regarding the inclusion of a study, team members will review disputed articles’ status until consensus is gained. Selected studies will then be assessed by at least two independent researchers for risk bias assessment using validated tools. Two researchers will pilot-test the data extraction form and independently screen the literature and extract the data. Data will be extracted and synthesised following a systematic review methodology and reported in accordance with the PRISMA guidelines to facilitate transparent reporting [1].

## Introduction

### Background

Cannabis for medicinal purposes originates from the plant *Cannabis sativa* and *Cannabis indica* and is composed of two main pharmacologically active compounds, cannabidiol (CBD), and tetrahydrocannabinol (THC). Cannabis used for medicinal purposes is referred to in a variety of ways, including medicinal cannabis, medical cannabis, cannabis as medicine, medical marijuana, pharmaceutical cannabis, prescribed cannabis, dispensed cannabis [2], cannabinoids and CBD oil. For the purposes of this review, we will use *cannabis for medicinal purposes* as a general term to denote all products including self-medicated cannabis, used for the treatment or relief of health-related symptoms.

Cannabis for medicinal purposes was first legalised in a number of jurisdictions in the United States in the 1990’s. These legislative changes, approving the use of cannabis for medicinal purposes under pre-specified conditions, were echoed across the globe with Israel and Canada in 2001, the Netherlands in 2003, Austria in 2008, Switzerland in 2011, Uruguay and Czechia in 2013, Croatia in 2015, Colombia and Australia in 2016, Germany in 2017, and Luxemburg, Portugal and the United Kingdom in 2018 [3-8]. Legislative conditions for the medical prescription of cannabis products for specified conditions are regulated by authorities such as the Australian Therapeutic Goods Administration (TGA), the United States Food and Drug Administration (FDA), and the European Medicines Agency (EMA). Whilst drug approval by a medical authority conveys safety and effectiveness for both patient and doctor, the prescription of cannabis for medicinal purposes under specified conditions potentially provides patients with the perception of relative safety. These regulatory changes have shown to increase the accessibility of cannabis for medicinal purposes for patients [9] leading to emerging preparations made available online and over the counter [10].

Despite only four cannabis products being approved for therapeutic use worldwide [11], the use of cannabis for medicinal purposes is on the rise internationally. While cannabis shows some levels of evidence for therapeutic benefits in the area of chronic pain, chemotherapy-induced nausea and vomiting, spasticity associated with multiple sclerosis and sleep disturbance [12], it is most commonly prescribed for the management of chronic pain and anxiety [13]. The use of cannabis for medicinal purposes during pregnancy is not recommended [14-19], yet, cannabis use in pregnancy is increasing [20-22]. Clinical evidence shows that cannabis use may be associated with adverse events such as low birth weight [19, 23-26], preterm birth [19] and childhood neurodevelopmental deficits [14, 19, 27, 28].

In addition, data collected from social media platforms regarding pregnancy and cannabis is showing three main online search trends: 1) safety and cannabis use during pregnancy; 2) the management of pregnancy-related symptoms including morning sickness, nausea, vomiting, headaches, pain, stress, and fatigue with cannabis use; and 3) cannabis use in the postpartum period [29]. This suggests an increased acceptance in the community.

The legalisation of cannabis for medicinal purposes, along with the prevalence of positive messages online and a lack of reporting of adverse events, may contribute to the perception that the use of cannabis for medicinal purposes carries little or no risk [22, 30]. Carr (2023) suggests that women consider that legalization leads to greater access and exposure to cannabis, increased acceptance of its use, and greater trust in cannabis retailers [22]. Additionally, there is a common belief in the safety of herbal medicines and a perception that plant-based medicines are safe during pregnancy [18].

### Evidence of impact of prenatal use of cannabis on the developing child

Evidence on the prenatal use of cannabis highlights potential negative impacts on the unborn child. These include low birth weight, reduced neonatal length, increased risk of admission in neonatal intensive care unit (NICU), smaller head circumference, earlier gestation age, and higher likelihood of preterm birth [31]. The most reported outcome for infants exposed to cannabis in utero was low birth weight [31]. One study found that cannabis use in early pregnancy and over a short period of exposure increased the risk of low birth weight and reduced head circumference [24]; however, 85% of the mothers in this study also reported prenatal use of tobacco, which is also linked to low birth weight and reduced head circumference [24]. Despite the potential effects of concomitant use of tobacco, three other studies on prenatal cannabis exposure reported decreased birth weight associated with the frequency of use [25, 26, 32], suggesting a possible dose response relationship.

Another reported outcome for infants exposed to cannabis in utero, compared to those non-exposed, is decreased gestational length [33, 34]. Additionally, a study comparing infant behaviour between newborns exposed to different levels of cannabis in utero and those not exposed found significant behavioural differences, such as decreased response to visual stimulus, inability to self-quiet and increased tremors and startles [35].

### Rationale

To date there is a lack of reviews that systematically investigate evidence of cannabis use for medicinal purposes in pregnancy. Specifically, this scoping review will map the available evidence of cannabis use for medicinal purposes for the management of the effects of pregnancy, including but not limited to hyperemesis gravidarum, nausea and vomiting, sleep disorder, morning sickness and restless legs syndrome; and for the ongoing management of pre-existing health conditions, and co-existing cannabinoids use disorders. In this review, evidence of the effects of prenatal cannabis exposure on newborns and infant development up to one year of age will also be identified.

To do this we will undertake a systematic scoping review of published research from 1996 to April 2024 to answer the following questions:

- What is the current evidence regarding the use of cannabis for medicinal purposes during pregnancy?
- What are the characteristics of the population using cannabis for medicinal purposes during pregnancy?
- What are the reported effects of medicinal prenatal cannabis use on neonates and infants up to 12 months old?

### Objectives

The objective of this review is to investigate the use of cannabis for medicinal purposes during pregnancy, focusing on the characteristics of the population using it, the current evidence supporting its use, and the reported effects on neonates and infants up to 12 months old.

## Method and Analysis

The review will be completed by November 2024 and will use the below PICOS framework:

|  |  |
| --- | --- |
| Population | Pregnant women above 18 years of age and below 40 years of age |
| Intervention | Using cannabis to treat pregnancy symptoms, other health conditions and cannabis addiction |
| Comparison | Usual care |
| Outcomes | • Evidence of cannabis use for medicinal purposes during pregnancy.<br>• Population characterisation of women using cannabis during pregnancy for medicinal purposes<br>• Evidence of the impact of cannabis use for medicinal purposes during pregnancy on foetus and up to 1 year postpartum |
| Studies | Reviews, randomised controlled trials, case–controlled, cross-sectional, and cohort studies |

The research questions will be addressed using a systematic scoping review approach. This scoping review protocol will be registered on Medrxiv and results will be reported according to the Preferred Reporting Items for Systematic Reviews and Meta-Analyses (PRISMA) statement [1].

### Search method

A comprehensive search will be conducted across multiple databases from 1996 to April 2024, including PubMed, MEDLINE, Embase and CINAHL. In addition, we will read through the list of references in each selected articles for other potential articles that may qualify for inclusion. Google Scholar will be used to search the citing articles of each study. The search criteria for PubMed can be found in appendix A. An identical keyword search will be applied to all four databases, the search will be translated for each database using Polyglot Search Translator [36].

### Study selection

Research relating to pregnant women who use cannabis for medicinal purposes and infants up to one year after birth who experienced in utero exposure to cannabis for medicinal purposes will be included in this review. Reviews, randomised controlled trials (RCTs), case-control, cross-sectional and cohort studies, that have been peer reviewed and published between 1996 and April 2024 as a research paper that report on the use of cannabis for medicinal purposes and its foetal, perinatal, and neonatal outcomes will be included. Due to the study population being pregnant women, for ethical reasons we do not expect the search to yield any RCTs, however, for completeness we have added this study type to the inclusion criteria.

The literature search will start from 1996, which corresponds to the year cannabis was first legalised for medical purposes in the United States (California). Only published studies in English and French will be included. The inclusion of the French language is intended to capture relevant literature from Canada and Europe. Excluded literature are editorials, letters, commentaries, conference papers, protocols, and book chapters. There will be no restriction on geographic location, however, restriction will be applied based on maternal age. Participants will need to be at least 18 years old for the treatment to be accessible on their own, and under 40 years of age to remove potential pregnancy complications associated with advanced maternal age. To better assign evidence to the use of cannabis for medicinal purposes, studies reporting on concomitant use of illicit substances (cocaine, methamphetamine, heroin, other), alcohol misuse and nicotine will be excluded when cannabis data cannot be isolated. In addition, studies or information related to cannabis preparations used solely for recreational purposes will be excluded from the analysis, as this study focuses on evidence of use of cannabis for medicinal purposes only. The study will also exclude evidence related to the use of synthetic cannabis due to its different pharmacological and epidemiological profiles [37].

All studies will be uploaded in Covidence [38] and screened based on titles and abstract as well as publication type. The full text articles that will be identified for inclusion will be reviewed independently and in duplicate by two researchers. AD will critique all articles, YB, RC, and CH will review a selection of articles, to ensure the literature is reviewed independently and in duplicate by at least two researchers. Where initial disagreement exists between reviewers regarding the inclusion of a study, team members will review the article’s status until consensus is gained.

### Study outcomes

Medicinal use will be identified from repeating and emergent themes based around the research question of this review: cannabis use for symptoms of pregnancy, cannabis as a medicine for specific health issues, impact of legislation on medicinal cannabis use, health professional discourse, and considerations of the mother.

Child outcomes are pre-defined based on previous studies, they may include preterm birth (<37 weeks), low birth weight (<2500g), small for gestational age, foetal length, birth weight, head circumference, gestation length, congenital anomalies, the need for intensive care unit placement, delay in growth, visual and speech milestones and changes in sleep patterns and behaviours. These outcomes are to be measured in utero, at birth, and up to one year of age.

## Data collection & Analysis

Two researchers will pilot-test the data extraction form and will independently screen the literature and extract the data. Refer to Appendix B for the data extraction form.

The following data will be collected for each article:

- Name of first author, year of publication, location.
- Study design, duration, sample size.
- Method of recruitment, study exclusion and inclusion criteria.
- Confounders, including reports of concomitant use of tobacco and alcohol, prescribed medications, vitamin supplements (e.g., folic acid), and potential bias associated with self-reporting.
- Characteristics of participants, including age, marital status, parity, ethnicity, income level, education level, and existing health conditions.
- Legislation status at the time of the study.
- Cannabis preparation type, method of delivery, and concentration.
- Reason for use and mode of access.
- Neonate/infant age at the time of the outcome measurement, and outcomes measured on the unborn child and up to twelve months after birth.

Risk assessment for bias will be performed by two independent researchers using current validated tools, selected according to each study design and the EQUATOR Network guidelines [39]. Risk bias assessment results will be reported using a traffic light diagram for each assessment. A green light will indicate that the study addressed the appraisal question in a clear acceptable way or presented low risk bias. An orange light will indicate that the study was unclear in addressing the appraisal question or presented some concerning risk bias. A red light will indicate that the appraisal question was not addressed in the study or presented high risk bias.

Given the anticipated heterogeneity of the data, we will categorise the reported outcomes into thematic groups identify by emerging themes or patterns. Results will be mapped by themes and present the type of study, methodology, findings, and limitations.

## Data Availability

All data produced in the present work are contained in the manuscript.

## Appendix A

### Search Criteria

The following databases will be searched from 1996 to April 2024: PubMed, MEDLINE/Ovid, Embase Classic+ Embase/Ovid and CINAHL Complete.

The below search describes the PubMed search strategy and will be translated for MEDLINE, Embase and CINAHL using Polyglot Search Translator.

Human only studies in English and French were applied in all four databases.

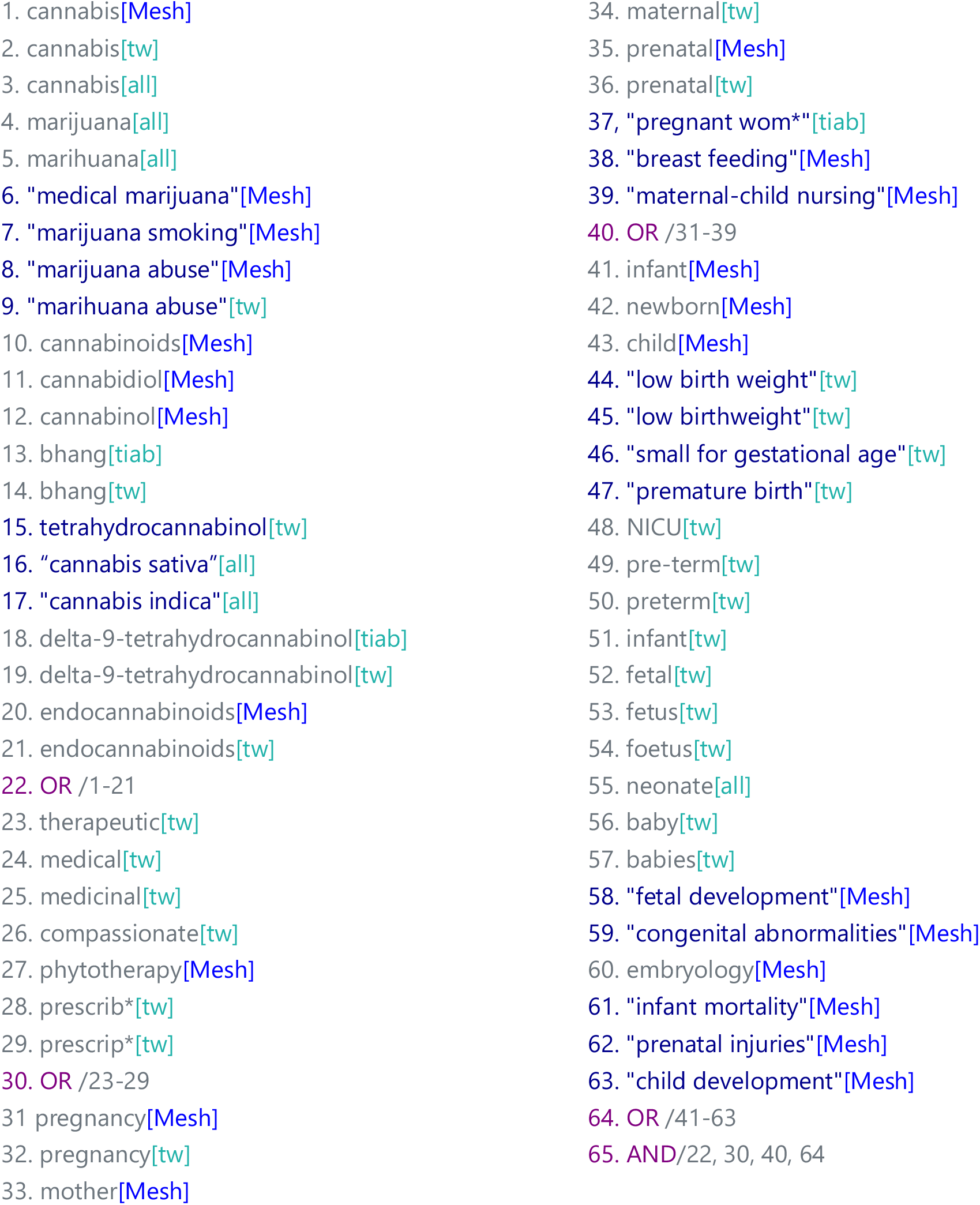

## Appendix B

### Data extraction form

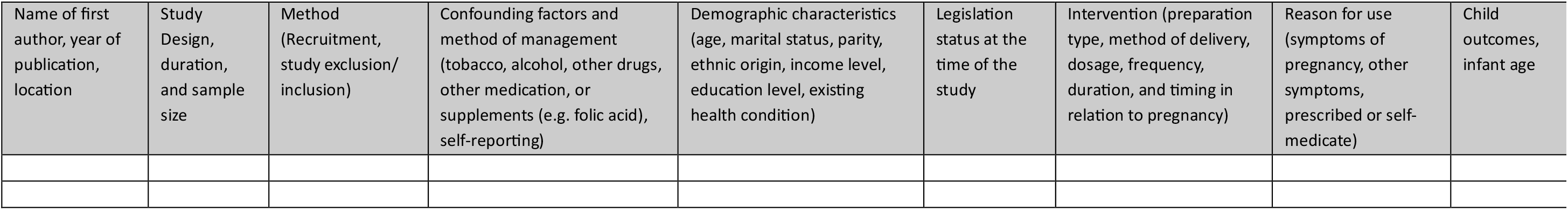

